# Situating Problematic Video Gaming and Psychotic-Like Experiences in the Adolescent Landscape of Affordances: A Cohort Study

**DOI:** 10.1101/2025.03.24.25324546

**Authors:** Vincent Paquin, Zoey Lavallee, Maxime Huot-Lavoie, Benson S. Ku, Covadonga M. Díaz-Caneja, Sinan Gülöksüz

## Abstract

**Background and aims:** Problematic gaming has been linked to increased levels of psychotic-like experiences (PLEs) in youth, but the role of environmental factors remains unclear. Using affordance theory, this study aimed to examine the association of problematic gaming with PLEs and the role of environmental factors.

**Methods:** Participants were 6492 youth (39.2% female) who reported playing video games, from the Adolescent Brain Cognitive Development Study in the U.S. Measures included problematic gaming, peer environment (number of close friends), school environment (teachers, activities, etc.), family environment (parental monitoring), and PLEs. We examined whether the peer, school, and family environments at age 12 were associated with problematic gaming and moderated its association with PLEs at age 13.

**Results:** Higher protective scores for the school and family environments at age 12 were independently associated with lower levels of problematic gaming at age 12 (respectively B=−0.15; 95% CI: −0.21, −0.10 and B=−2.39; 95% CI: −2.71, −2.07) and age 13 (B=−0.05; 95% CI: −0.10, −0.00 and B=−0.79; 95% CI: −1.11, −0.47). The peer environment was not associated with problematic gaming. Higher levels of problematic gaming at age 12 were associated with higher levels of PLEs at age 13 (B=0.13; 95% CI: 0.09, 0.17), with no significant interaction with the environmental variables.

**Discussion and conclusions:** Positive school and family environments may be protective against problematic gaming in adolescence but do not appear to attenuate the putative effect of problematic gaming on PLEs. The results provide partial support to an affordance-based conceptualization of problematic gaming.

## INTRODUCTION

Psychotic-like experiences (PLEs) are unusual thoughts and perceptions similar to the symptoms of psychotic disorders, such as paranoia and hallucinations, but of lesser intensity. PLEs are found in 9-14% of adolescents (Healy et al., 2019; Lindgren & Therman, 2024) and are associated with a greater risk of psychotic, mood, and anxiety disorders (McGrath et al., 2016; Sullivan et al., 2020).

A potential risk factor for PLEs is problematic gaming, which is commonly defined following the components model of addiction : salience (attributing importance to gaming), tolerance (playing more and more), mood modification (using games to feel better), relapse (losing control of the amount of gaming), withdrawal (having negative experiences from discontinuing gaming), and conflict (having functional impairment or distress related to gaming) (Billieux, Flayelle, Rumpf, & Stein, 2019; Griffiths, 2005). Problematic gaming should therefore be distinguished from merely playing games, which is not in itself a risk factor for PLEs (Paquin, Ferrari, et al., 2024; Paquin, Philippe, et al., 2024) and which can provide experiences that are beneficial for mental health, such as social connections, identity development, and stress relief (Ballou, Hakman, Vuorre, Magnusson, & Przybylski, 2024). There is interest in improving the management of problematic gaming in youth, including to mitigate its potential impact on the clinical trajectories of psychotic disorders (Huot-Lavoie et al., 2024). Problematic gaming was positively correlated to PLEs in a cross-sectional study (Zhang et al., 2022) and was suggested to contribute to psychotic symptoms in clinical case reports (Huot-Lavoie et al., 2022). One explanation for the potential effect of problematic gaming on PLEs is its association with increased social withdrawal, a risk factor for PLEs (Narita et al., 2024).

There is a need, however, to accurately differentiate problematic from non-problematic gaming. Some of the typical items used to measure problematic gaming, such as “frequently thinking about games”, are thought to lack specificity by tapping into a harmless proclivity for games (Fournier et al., 2023; Wichstrøm, Stenseng, Belsky, von Soest, & Hygen, 2019). This is why, in contrast to survey-based assessments of problematic gaming, the clinical diagnosis of gaming disorder requires the presence of functional impairment or psychological distress (King, Billieux, Behm, & Delfabbro, 2025). For cohort studies that include only survey-based assessments of problematic gaming, the integration of environmental factors may help distinguish problematic from non-problematic gaming.

### Affordance theory of problematic gaming

Environmental factors in problematic gaming can be understood using affordance theory (Glackin, Roberts, & Krueger, 2021; Lavallee & Osler, 2024), which posits addictive behaviour as not merely the result of individual psychological features but as arising from and being sustained by features of the person’s environment (Lavallee & Osler, 2024, p. 380). Affordances are possibilities of action or experience, including emotional experience, that are enabled by the environment: a pen affords writing, close friendships afford intimacy, and games afford enjoyment (Gibson, 1979; Rietveld & Kiverstein, 2014). Affordances are not purely external features of the world or internal mental states: they are constituted by both the features of the environment and the embodied needs and capabilities of the person (Kiverstein, 2020). For instance, a game might afford the possibility of being played to two people, but if the former person is a passionate gamer and the latter dislikes games, then its affordance of play will only be alluring for the former person.

For Lavallee & Osler (2024), addiction is characterized by the narrowing of affordances to those related to the addictive behavior. Accordingly, in its problematic form, gaming starts as a central means for someone to fulfill their psychological needs (e.g., experiencing social connection, autonomy, competence) (Allen & Anderson, 2018) and becomes pathological as the person reorganizes their life to pursue gaming, to the detriment of nurturing other spheres of their social environment that could enable alternative affordances (Billieux et al., 2017; Narita et al., 2024). In contrast, environments that provide a wider range of possibilities for the fulfillment of psychological needs might help protect against a monopolizing effect of gaming-related affordances. Further, a person’s maintenance of protective affordances across their social environment might be a sign that their gaming behavior has not evolved into an addiction. Altogether, this affordance-based understanding reiterates the clinical recognition of functional impairment as an essential feature of gaming disorder (Billieux et al., 2017) and expands on it by formalizing the relationship between addiction and the person’s environment.

The implication of affordance theory is that we can better evaluate problematic gaming and its association with PLEs by integrating information about a person’s perceived affordances in their environment. During adolescence, the peer, school and family environments are important determinants of mental health (McGorry et al., 2024). Experiences of hostility or exclusion in these spheres are associated with greater engagement in video games and higher levels of PLEs (Paquin, Ferrari, et al., 2024). The presence of protective affordances enabled by peer, school, and family environments might be a sign that an adolescent’s gaming is not problematic, and it could attenuate the impact of problematic gaming on PLEs by mitigating the exposure to hostility, exclusion, or other risk factors for PLEs. In sum, higher survey scores for problematic gaming may be less reliable markers of PLE risk among adolescents who report better protective features in their peer, school, or family environment.

This study aimed to examine, through the lens of affordance theory, the prospective association of problematic video gaming during adolescence with PLEs. To this end, we used a population-based cohort to evaluate the role of protective affordances in the peer, school, and family environments in this association. We hypothesized that the protective affordances indexed by environmental variables would be associated with lower levels of problematic video gaming and that they would attenuate the association of problematic gaming with subsequent PLEs.

## METHODS

### Participants

Data was drawn from the Version 5.1 of the Adolescent Brain and Cognitive Development (ABCD) Study (Garavan et al., 2018), which includes visits between September 1^st^, 2016 and January 15, 2022. The ABCD Study is a population-based cohort of 11,868 children aged 9-10 years at baseline and followed annually. Participants were recruited from randomly selected schools within a 50-mile radius of 22 research sites across the United States. The present analyses focused on follow-up data at years 2 and 3 (ages 12-13). We chose year 2 because it is the first time point where problematic gaming was measured, and we examined PLEs in the next year to estimate a prospective association. Analyses were restricted to participants with complete data on analytic variables (n=8886). Because the problematic gaming questionnaire was not administered to non-gaming participants, the analyses were further restricted to participants who reported playing video games at year 2, leading to a final sample of n=6492 (see Results for participant characteristics). To index affordances in the peer, school and family environments, we used self-reported rather than informant-reported measures, following the notion that affordances reflect not only the features of the environment but also the person’s relation to it, including their perception (Lavallee & Osler, 2024).

### Measures

PLEs were measured at years 2 and 3 using the Prodromal Questionnaire–Brief Child Version (PQBC), which includes 21 items on the occurrence of PLEs in the past month over a 6-point scale: 0 = no distress, 1 = yes but low distress, and 2–6 = yes with increasing distress. The questionnaire was read to participants by research assistants. The PQBC in the ABCD shows good evidence of construct validity as a measure of the psychosis spectrum (Karcher et al., 2018). We used the total distress scores (range: 0– 126), on which we applied a log-transformation to attenuate their skewed distribution as done previously (Karcher et al., 2018, 2022), followed by a multiplication by 10 to reduce scaling differences relative to other variables.

Problematic gaming was measured at years 2 and 3 with the Video Game Addiction Questionnaire. This scale was adapted from the widely used Bergen Facebook Addiction Scale (Andreassen, TorbjØrn, Brunborg, & Pallesen, 2012) and includes 6 items based on the components model of addiction, such as “I spend a lot of time thinking about playing video games” and “I play video games so much that it has had a bad effect on my schoolwork or job” (Bagot et al., 2022). Items are rated on a scale of 1 = “Never” to 6 = “Very often” and summed (range: 6-36).

Peer environment was measured at year 2 as the number of close friends, which was calculated as the sum of 3 items about close friends of female, male, or other gender. We chose this measure as the proxy for peer environment based on a previous study that found that the number of close friends attenuated the association of neighborhood fragmentation with PLEs in the same cohort (Ku, Ren, et al., 2024). We preferred this measure over other peer-related variables from the ABCD Study, which assessed risk rather than protective factors (e.g., bullying) or targeted specific behaviors not related to the present research question (e.g., peer encouragement for not using drugs). Given the wide distribution of the variable (median: 5; range: 0-200), we applied winsorizing to limit the influence of outlier values, setting values above the 95^th^ percentile to the 95^th^ percentile value, which was 17.

School environment was measured at year 2 with the School Environment subscale of the School Risk and Protective Factor Survey (Arthur et al., 2007; Hamilton et al., 2011). This subscale includes 6 items addressing protective aspects of the school environment, such as “In my school, students have lots of chances to help decide things like class activities and rules” and “I get along with my teachers”. The subscale score (range: 6-24) is the sum of each item rated on scale of 1 = “NO!” to 4 = “YES!”. This measure was previously found to moderate the association between neighborhood-level deprivation and hippocampal volumes in the ABCD (Ku, Aberizk, et al., 2024).

Family environment was measured at year 2 using the Parental Monitoring Survey, which was adapted for the ABCD Study from two other measures (Karoly, Callahan, Schmiege, & Ewing, 2016; Stattin & Kerr, 2000). The questionnaire includes 5 items assessing a parent’s active care efforts regarding the whereabouts of the child, such as “How often do your parents/guardians know where you are?” and “If you are at home when your parents or guardians are not, how often do you know how to get in touch with them?”. Items are rated on a scale of 1 = “Never” to 5 = “Always or almost always” and averaged (range: 1-5).

Other covariates were reported by caregivers at baseline: age, sex assigned at birth, higher parental education (defined as at least one of the parents or caregivers having a bachelor’s degree), income-to-needs ratio, ethnoracial group (Asian, Black, Hispanic, White, and other), and family history of psychosis. Income-to-needs ratio was calculated as the midpoint of the income category divided by the federal poverty line for the respective household size (Rakesh, Zalesky, & Whittle, 2022). Family history of psychosis was assessed as any vs. none across first- and second-degree relatives using the Family History Assessment Module Screener (van Dijk, Murphy, Posner, Talati, & Weissman, 2021).

### Statistical analyses

We conducted the analyses in R version 4.3.3 between February and March 2025 following our preregistered study protocol (https://osf.io/f53ah/) and published the analytic codes online (https://osf.io/4zfxd/). Deviations from the protocol were the inclusion of the ethnoracial group as a covariable and the addition of sensitivity analyses to confirm the robustness of the findings (Supplementary Table 1). From the total ABCD sample, characteristics of participants with and without missing data on analytic variables were compared to identify meaningful differences, using effect sizes ≥ 0.100 rather than P-values (Vandenbroucke et al., 2007). Participants who did not report playing games at year 2 were removed after to arrive at the final sample.

For subsequent analyses, nominal statistical significance was set at P-value < .05. First, we used a linear mixed model to examine the associations of the peer, school, and family environments at year 2 with levels of problematic gaming at year 2. The model had random intercepts for sites and families to account for clustered observations. Covariates were age, sex, parental education, income-to-needs ratio, ethnoracial group, family history of psychosis at baseline, and PLEs at 2 years. Second, we used another linear mixed model to examine the association between levels of problematic gaming at year 2 and PLEs at year 3. Covariates were the same as above, including the peer, school, and family environment variables. Third, we examined whether the peer, school, and family environment variables at year 2 moderated the association between problematic gaming at year 2 and PLEs at year 3. We modeled 2-way interactions for each pair of environmental variable and problematic gaming in three separate models. P-values of interaction terms were adjusted for false discovery rate using the Benjamini-Hochberg method (Benjamini & Hochberg, 1995).

As sensitivity analyses, we tested a 2-factor model of problematic gaming using confirmatory factor analysis. We drew from recent evidence of two latent factors (“engagement”, items 1-2; and “addiction”, items 3-6) in a similar 6-item scale of problematic social media use (Fournier et al., 2023). Following the analytic code provided by Fournier et al. (2023), we applied the weighted least squares means and variance adjusted robust estimation method, and we assessed the quality of the model using conventional fit indices. Furthermore, we also examined whether the peer, school, and family environment variables at year 2 were prospectively associated with problematic gaming at year 3. Data on problematic gaming at year 3 were available for 5866 (2052 [35.0%] female) participants.

### Ethics

The study procedures were carried out in accordance with the Declaration of Helsinki. Children provided verbal assent and caregivers provided written informed consent to the ABCD research protocol, which was approved by the Institutional Review Board at each participating site: https://abcdstudy.org/consortium_ members/.

## RESULTS

Relative to participants excluded from analyses because of missing data, the sample of included participants featured greater proportions of higher parental education and White participants, smaller proportions of Asian, Black, and Hispanic participants, and a higher income-to-needs ratio on average (effect sizes ≥ 0.100; Table 1). Bivariate correlations between all analytic variables are in Figure 1.

**Figure 1.**
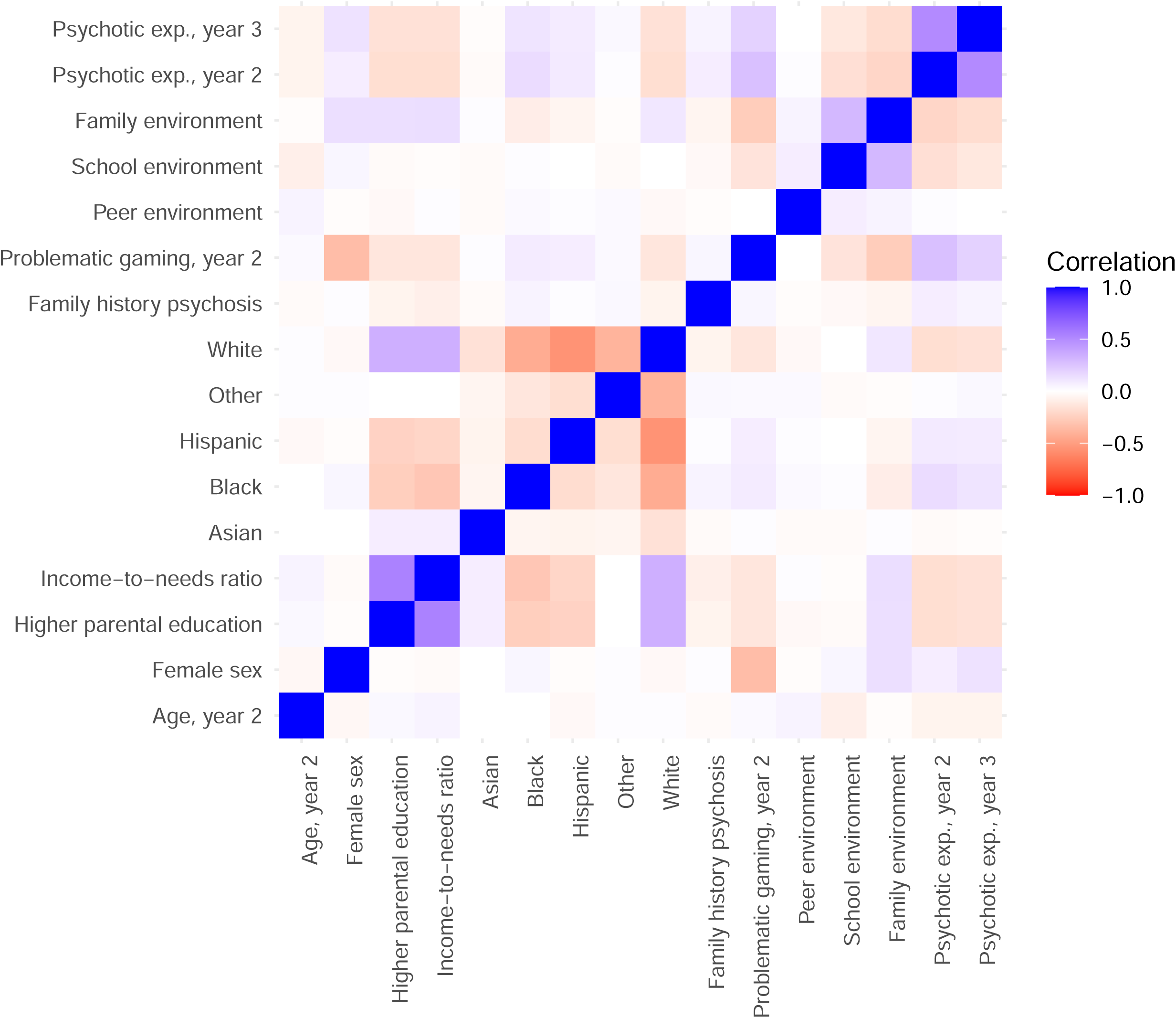
Bivariate correlations between variables of the main analyses. Correlations are Pearson’s r. For each ethnoracial group, the comparator is all other ethnoracial groups combined. Scores for psychotic-like experiences were log- transformed.

**Table 1.**
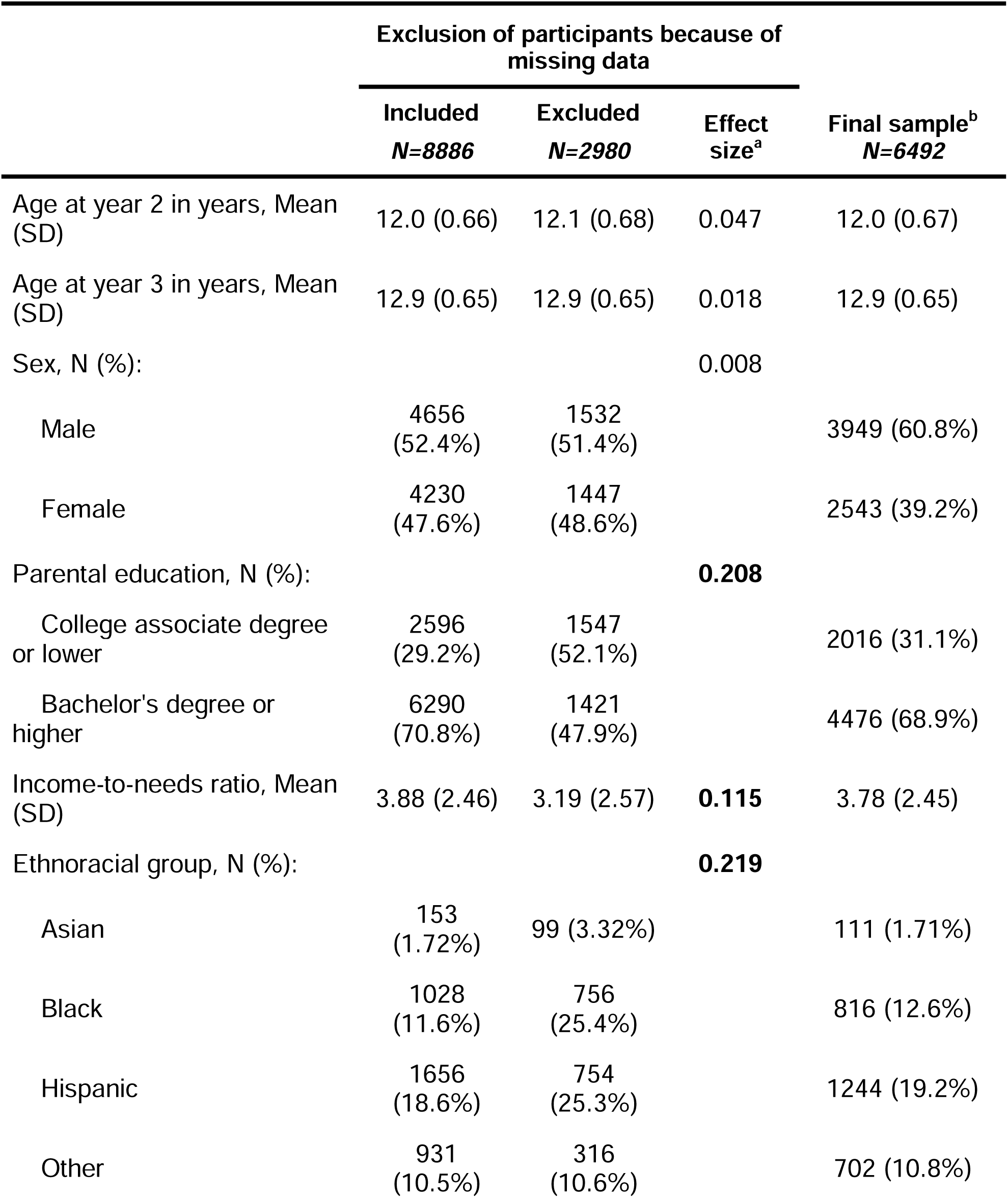
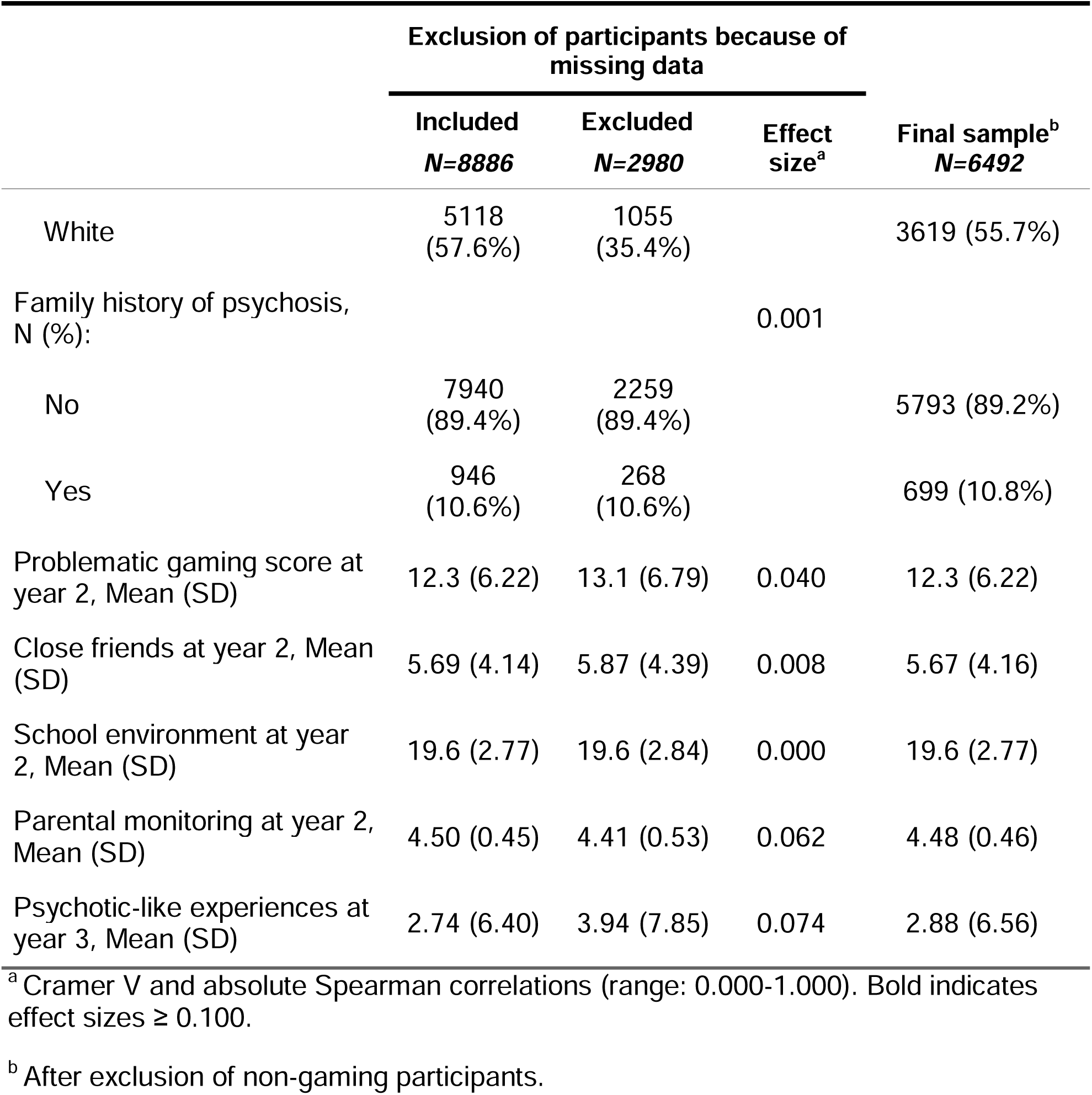
Characteristics of participants included in and excluded from the analyses.

### Main analyses

Higher scores for protective school and family environments were concurrently associated with lower levels of problematic gaming at year 2, whereas the peer environment variable was not significantly associated with problematic gaming in the same year (Figure 2, top left). Higher levels of problematic gaming at year 2 were significantly associated with higher levels of PLEs at year 3 (Figure 2, bottom left). There was a significant interaction effect between school environment and problematic gaming at year 2 after correction for false discovery rate (Figure 2, bottom right). Probing the interaction with school environment indicated that at higher levels of protective school environment, there was a *larger* association between higher levels of problematic gaming at year 2 and higher levels of PLEs at year 3. However, the estimated trends did not significantly differ over the lowest and upper quartiles of the school environment scores (Supplementary Figure 1). This means that any significant interactive effect would be confined to very low or very high values of school environment scores. Other interactions of problematic gaming with the peer and family environment variables were not significant.

**Figure 2.**
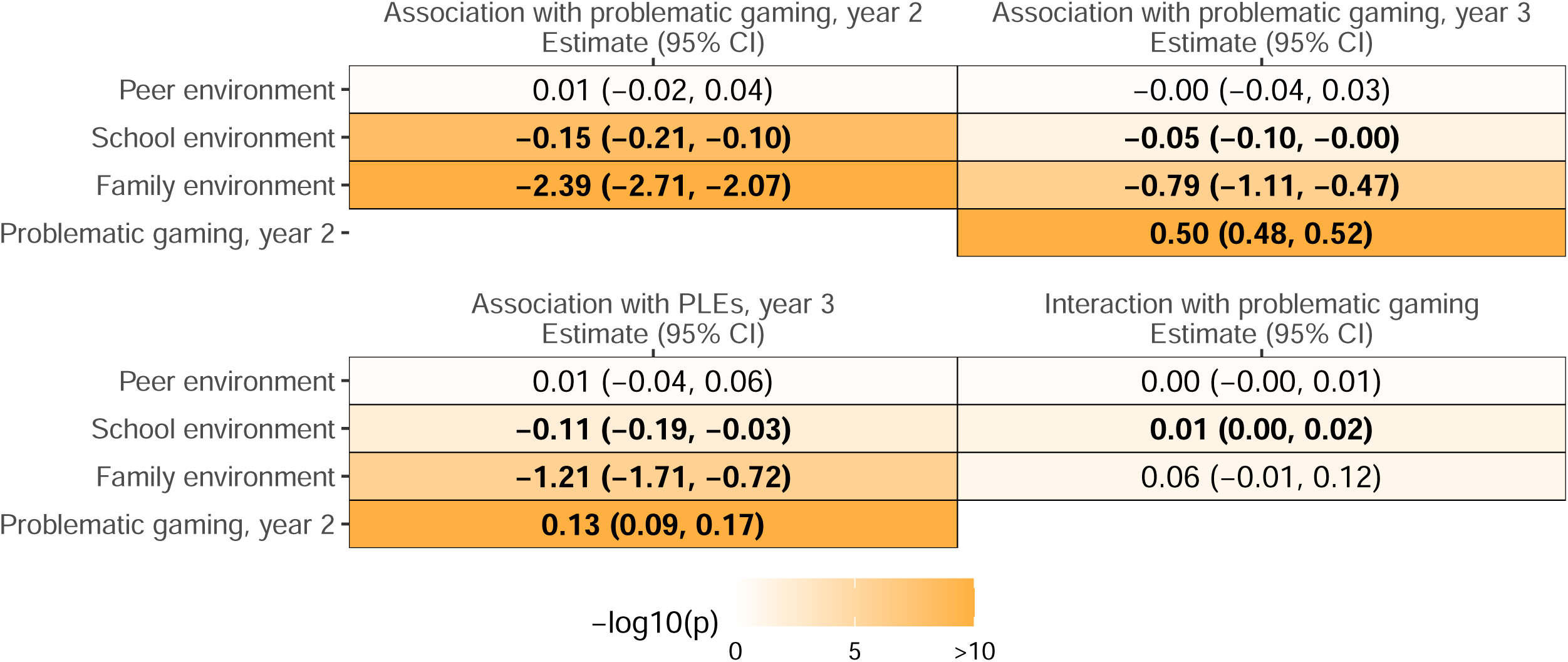
Adjusted associations between problematic gaming, environmental factors and psychotic-like experiences. Bold indicates p<.05. All models were adjusted for age, sex, parental education, income-to-needs ratio, ethnoracial groups, and family history of psychosis. Psychotic- like experiences (PLEs) at year 3 was further adjusted for PLEs at year 2 (both log- transformed and rescaled), and problematic gaming at year 3 was adjusted for problematic gaming at year 2. For interactions with problematic gaming (bottom right), the outcome is PLEs at year 3.

### Sensitivity analyses

A two-factor model of problematic gaming at year 2 provided a good fit to the data, but the two factors were almost perfectly correlated, preventing their evaluation as separate predictors of PLEs (Figure 3). Higher scores for protective school and family environments at year 2 were significantly associated with lower levels of problematic gaming at year 3 (Figure 2, upper left).

**Figure 3.**
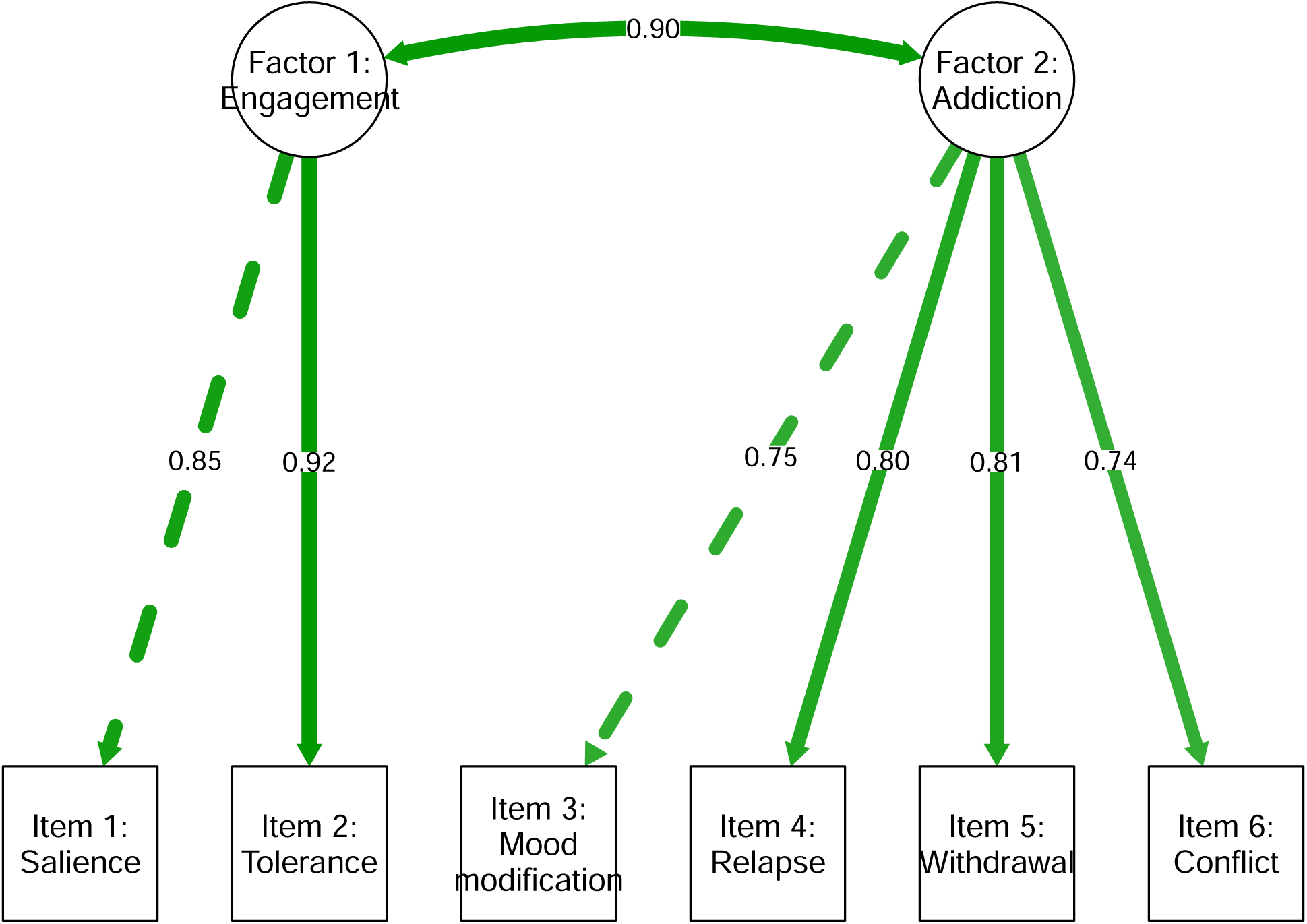
Confirmatory factor analysis of a 2-factor model of problematic gaming at year 2. Model based on Fournier et al. (2023). Fit indices: χ^2^(8)=136.71, p<.001; root mean square error of approximation (RMSEA)=0.05; comparative fit index (CFI)=0.997, Tucker-Lewis index (TLI)=0.995.

## DISCUSSION AND CONCLUSIONS

In this cohort of early adolescents, problematic video gaming was prospectively associated with higher levels of PLEs, independently of prior PLEs and other sociodemographic and environmental factors. This association is consistent with a previous cross-sectional study on problematic gaming and PLEs in adolescents (Zhang et al., 2022) and longitudinal studies that examined problematic internet use, a concept similar to problematic gaming, in association with PLEs in adolescents (Narita et al., 2024) and college students (Peng & Zou, 2025). Although this association is observational and subject to unmeasured confounding influences, it could reflect a causal effect of problematic gaming on the emergence of PLEs as a result of increased social isolation, interpersonal conflicts, or other socio-environmental disruptions (Narita et al., 2024; Paquin, Ferrari, Sekhon, & Rej, 2023).

The results mostly supported our first hypothesis, which was that protective affordances indexed by the peer, school, and family environment variables would be associated with lower levels of problematic gaming. We found such protective associations for the school and family environment variables across problematic gaming at years 2 and 3. In line with these results, systematic reviews support the role of school and parental factors in the risk of problematic gaming during adolescence (Coşa, Dobrean, Georgescu, & Păsărelu, 2023; Gao, Wang, & Dong, 2022). For example, in a cohort of 833 youth aged 11-14 years in China, greater parent-adolescent closeness and school connectedness were prospectively associated with lower levels of problematic gaming (Zhu, Zhang, Yu, & Bao, 2015). However, we found no association of the peer environment variable (close friends) with problematic gaming, contrary to previous studies in which having fewer closer friends was cross-sectionally associated with higher levels of problematic gaming in young adults in Turkey (Ünal, Gökler, & Turan, 2022) and the U.S. (Ohayon & Roberts, 2021), as well as in children in Japan (Yamada, Sekine, & Tatsuse, 2023). We chose this indicator as the “best” indicator available in the ABCD dataset, in our opinion, to assess the protective peer environment, but admittedly it has at least two limitations: it focuses on the quantity rather than the quality of friendships, and it does not capture whether the friends encourage or discourage problematic gaming behaviors. These two limitations may explain the null association with problematic gaming.

The results did not support our second hypothesis that higher scores for protective peer, school, and family environments would attenuate the association between problematic gaming and PLEs. The only significant interaction was with the school environment, but its direction was opposite to what we hypothesized (i.e., a better school environment seemingly amplified the association between problematic gaming and PLEs) and too weak to produce significant differences within the lower and upper quartiles of school environment scores. Overall, we can conclude that the prospective risk of PLEs that is putatively conferred by problematic gaming remains stable across a range of peer, school, and family environmental characteristics.

In addition, the measure of problematic gaming did not lend itself to a two-factor model, despite previous arguments that sum scores on this type of scale conflates indicators of engagement (e.g., frequently thinking about games) with indicators of addiction (e.g., functional impairment) (Fournier et al., 2023). The two-factor model is another proposal to better differentiate problematic from non-problematic gaming, following the notion that addiction to games is characterized by functional impairment or psychological distress (King et al., 2025). The present study provides initial evidence that these two approaches– interaction and factor analyses – do not help discriminate the contribution of gaming to the emergence of PLEs, beyond what the sum score of problematic gaming already indicates, and within the limits of the environmental measures examined here.

Limitations of the study include the reliance on empirical proxies that incompletely index the concepts that we sought to investigate, such as affordances in the peer, school, and family environments. Extended assessments using more detailed measures of the environment, ambulatory data, or qualitative inquiries could address this gap. We examined the two timepoints (years 2 and 3) of the ABCD where data on problematic gaming was available, but shorter timeframes would elucidate more immediate interdependencies between gaming and the environment, whereas longer timeframes would be necessary to discriminate between transient and persistent PLEs. Problematic gaming was assessed through self-report in a community-based sample, limiting the transferability of the results to clinical populations where functional impairment and distress are subject to a personalized evaluation by the clinician. External validity may also be limited by the effect of attrition, notably as a function of educational attainment, household income, or ethnoracial groups, which were identified as meaningful differences between participants with and without missing data.

### Implications for an affordance theory of problematic gaming

To our knowledge, this is the first conceptualization of problematic gaming that is explicitly grounded in an affordance theory of addiction (Lavallee & Osler, 2024). We defined problematic gaming as the concentrated pursuit of affordances related to games to the detriment of pursuing other affordances in the person’s environment. Affordance theory emphasizes that problematic gaming depends not only on the player’s psychological features and behaviors, or the features of the game, but also on the broader environment in which the player and game are situated. Other accounts of problematic gaming have similarly acknowledged the role of the environment in the emergence or maintenance of problematic gaming, with some divergences on the relative importance of individual vs. social dimensions, as well as whether “addiction” is an appropriate term (e.g., Kardefelt-Winther, 2017; Király, Koncz, Griffiths, & Demetrovics, 2023; Snodgrass et al., 2014).

Our contention is not that affordance theory is superior to other explanations, but rather that it has some utility in understanding the process and mental health ramifications of problematic gaming. An affordance-based conception calls for a close examination of what affordances, in the person’s environment, protect against or suffer from the emergence of problematic gaming. The present results suggest that protective affordances in the school and family environments may protect against problematic gaming or at least may be indirect markers of non-problematic gaming. Of interest for future research, an affordance-based conception of problematic gaming can integrate more granular theories of the psychological motivations for gaming, such as self-determination theory and escapism (Allen & Anderson, 2018; Giardina et al., 2024), as well as the role of game characteristics in promoting addiction (Flayelle et al., 2023). By rethinking problematic gaming in terms of affordances in and out of games, researchers and clinicians may find affordance theory a helpful device for holding together the psychological, environmental, and game-related factors that shape problematic gaming.

## FUNDING SOURCES

VP is supported by an award from the Fonds de recherche du Québec – Santé and Ministère de la santé et des services sociaux (FRQS-MSSS). BSK received support from the National Institute of Mental Health (NIMH K23-MH129684). SG and CDC received support for this work as part of the Youth-GEMs project, funded by the European Union’s Horizon Europe Program under Grant Agreement Number 101057182. CDC received support from Instituto de Salud Carlos III, Spanish Ministry of Science and Innovation (PI20/00721, PI23/00625) and the European Commission (HE YOUTHreach 101156514). Data used in the preparation of this article were obtained from the Adolescent Brain Cognitive Development (ABCD) Study (https://abcdstudy.org), held in the NIMH Data Archive (NDA). The ABCD Study® is supported by the National Institutes of Health and additional federal partners under award numbers U01DA041048, U01DA050989, U01DA051016, U01DA041022, U01DA051018, U01DA051037, U01DA050987, U01DA041174, U01DA041106, U01DA041117, U01DA041028, U01DA041134, U01DA050988, U01DA051039, U01DA041156, U01DA041025, U01DA041120, U01DA051038, U01DA041148, U01DA041093, U01DA041089, U24DA041123, U24DA041147. A full list of supporters is available at https://abcdstudy.org/federal-partners.html. A listing of participating sites and study investigators can be found at https://abcdstudy.org/consortium_members/. This manuscript reflects the views of the authors and may not reflect the opinions or views of the NIH or ABCD consortium investigators. The ABCD data used in this report came from http://dx.doi.org/10.15154/z563-zd24.

## AUTHORS’ CONTRIBUTION

VP designed the present study (a secondary analysis of the ABCD), conducted the analyses and drafted the manuscript. SG provided supervision. VP, SG, ZL, MHL, BSK, and CDC contributed to revising the study protocol, interpreting the results and revising the manuscript. VP had full access to all data in the study and takes responsibility for the integrity of the data and the accuracy of the data analysis. ABCD consortium investigators designed and implemented the ABCD study or provided data but did not participate in the analysis or writing of this report.

## DISCLOSURES

CDC has received honoraria and/or travel support from Angelini, Johnson&Johnson, and Viatris for work unrelated to this study.

## Data Availability

Data used in the preparation of this article were obtained from the Adolescent Brain Cognitive Development (ABCD) Study (https://abcdstudy.org), held in the NIMH Data Archive (NDA). The ABCD data used in this report came from https://nda.nih.gov/study.html?id=2313.

## Notes

### Author Declarations

Data used in the preparation of this article were obtained from the Adolescent Brain Cognitive Development (ABCD) Study (https://abcdstudy.org), held in the NIMH Data Archive (NDA). The ABCD Study is supported by the National Institutes of Health and additional federal partners under award numbers U01DA041048, U01DA050989, U01DA051016, U01DA041022, U01DA051018, U01DA051037, U01DA050987, U01DA041174, U01DA041106, U01DA041117, U01DA041028, U01DA041134, U01DA050988, U01DA051039, U01DA041156, U01DA041025, U01DA041120, U01DA051038, U01DA041148, U01DA041093, U01DA041089, U24DA041123, U24DA041147. A full list of supporters is available at https://abcdstudy.org/federal-partners.html. A listing of participating sites and study investigators can be found at https://abcdstudy.org/consortium_members/. This manuscript reflects the views of the authors and may not reflect the opinions or views of the NIH or ABCD consortium investigators. The ABCD data used in this report came from http://dx.doi.org/10.15154/z563-zd24.

