## Supplementary for "Situating Problematic Video Gaming and Psychotic-Like Experiences in the Adolescent Landscape of Affordances: A Cohort Study"

**Supplementary Material**

### Supplementary Table 1. Deviations from the preregistered study protocol

| **Deviations** | **Explanation** |
| --- | --- |
| We used the ABCD Data Release Version 5.1 instead of 6.0. | The release of 6.0 was delayed, and Version 5.1 contained the required time points for the planned analyses. |
| We included the ethnoracial group as a covariate, in addition to other covariates already specified in the protocol. | We considered that the ethnoracial group might confound the association between problematic video gaming and psychotic-like experiences. |
| We conducted an additional (sensitivity) analysis to examine the association of protective environmental factors at year 2 with problematic gaming at year 3. | Given the associations between two of these environmental variables (school and family) with concurrent problematic gaming at year 2, we were interested in examining whether the same associations would hold prospectively with problematic gaming at year 3. |
| We initially planned to report the association between problematic gaming at year 2 and psychotic-like experiences (PLEs) at year 3 following 2 sets of covariates:   - Baseline covariates - Baseline covariates, PLEs at 2 years, and the peer, school, and family environmental variables.   In the article, we only report the second set that comprises all variables. | Because the results were consistent across the two sets of variables, we opted not to report the first one (coefficient=0.361; 95% CI: 0.321, 0.401) to reduce the word count. Also, the omission of PLEs at year 2 in the first set of covariates was an error in the protocol. |

The preregistered study protocol is available on the Open Science Framework: <https://osf.io/f53ah>

### Supplementary Figure 1. Interaction between problematic gaming and the school environment at year 2

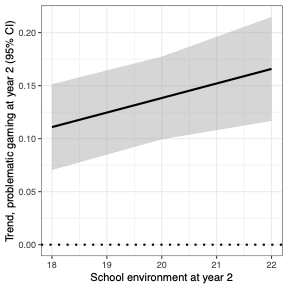

The outcome is psychotic-like experiences (PLEs) at year 3. The interaction was probed using estimated marginal trends (*emmeans* package in R). The magnitude of the association between gaming and PLEs (Y-axis) increases with higher scores for protective school environment (X-axis). However, as the overlapping confidence intervals (shaded area) demonstrate, the apparent increase in the strength of association is not statistically significant.
